# A Randomized Controlled Double-Blinded Split-Face Prospective Clinical Trial to Assess the Efficacy, Safety, and Tolerability of a Novel 3-Step Routine Compared to Benzyl Peroxide for the Treatment of Mild to Moderate Acne Vulgaris

**DOI:** 10.1101/2023.11.12.23298113

**Authors:** Alison Gern, Jessica Walter, Shuai Xu, Paras P. Vakharia

**Affiliations:** Department of Dermatology - Northwestern University Feinberg School of Medicine (Chicago, IL); Department of Obstetrics and Gynecology – Northwestern University Feinberg School of Medicine (Chicago, IL); Department of Pediatrics (Dermatology) - Northwestern University Feinberg School of Medicine (Chicago, IL); Sibel Health Inc. (Chicago, IL)

## Abstract

Adult acne vulgaris affects up to 43-51% of individuals. While there are numerous treatment options for acne including topical, oral, and energy-based approaches, benzoyl peroxide (BPO) is a popular over the counter (OTC) treatment. Although BPO monotherapy has a long history of efficacy and safety, it suffers from several disadvantages, most notably, skin irritation, particularly for treatment naïve patients. In this prospective, randomized, controlled, split-face study, we evaluated the comparative efficacy, safety, and tolerability of a novel 3-step azelaic acid, salicylic acid and graduated retinol regimen (Geologie Clear System) versus a common OTC BPO-based regimen (Proactiv Solution) over 12 weeks. A total of 37 adult subjects with self-reported mild to moderate acne vulgaris were recruited. A total of 21 subjects underwent a 2-week washout period and completed the full study with 3 dropping out due to product irritation from the BPO routine, and 13 being lost to follow-up. Detailed tolerability surveys were conducted at Week 4. Additional surveys on tolerability and product preferences were collected monthly, at Week 4, Week 8, and Week 12. A blinded board-certified dermatologist objectively scored the presence and type of acne lesions (open or closed comedones, papules, pustules, nodules, and cysts) at baseline, Week 4, Week 8, and Week 12. Patients photographed themselves and uploaded the images using personal mobile phones. Detailed Week 4 survey results showed across 25 domains of user-assessed product performance, Geologie outperformed the BPO routine in 19 (76%) which included domains in preference (e.g. “I would use this in the future), performance (“my skin improved” and “helped my acne clear up faster”). Geologie users reported less facial redness, itching, and burning, though differences did not reach statistical significance. In terms of efficacy, both products performed similarly, reducing total acne lesions by 36% (Geologie) and 40% (BPO routine) by Week 12. Overall, accounting for user preferences and tolerability Geologie was more preferred than the BPO routine in 79% of domains (22/28). Differences in objective acne lesion reduction were not statistically significant (p=0.97). In a randomized split-face study, a 3-step azelaic acid, salicylic acid, and graduated retinol regimen delivered similar acne lesion reduction, fewer user dropouts, greater user tolerability, and higher use preference compared to a 3-step BPO routine based in a cohort of participants with mild-to-moderate acne vulgaris.

## Introduction

Acne vulgaris is one of the most common human diseases in adults affecting 43-51% of individuals between the ages of 20 to 29 and up to 35% of individuals between the ages of 30 to 39.^1^ The disease can be disfiguring with a profound psychological impact, contributing to both anxiety and depression.^2^ Currently, there are a wide range of topical, oral, and energy-based therapies for acne. Topical, over-the-counter (OTC) therapies are popular, representing a global $5 billion USD market.^3^ Within this category of topical treatments, benzoyl-peroxide (BPO) based therapies remain one of the most commonly used worldwide.

While BPO as a monotherapy is an effective treatment for acne,^4^ it has significant side effects that affect adherence, including skin irritation and patient intolerability, particularly among individuals with sensitive skin. In one study, 35% of BPO users noted side effects with a discontinuation rate of 44% at 6 months.^5^ Importantly, the prevalence of sensitive skin in the adult population is greater than 70% in a recent systematic review.^6^ Thus, new OTC regimens offering comparable efficacy with greater tolerability compared to existing BPO therapies would be beneficial to adult patients with mild-to-moderate acne vulgaris.

We hypothesized a novel 3-step regimen with 3 primary anti-acne ingredients (azelaic acid, salicylic acid, and retinol) would offer similar efficacy and greater tolerability compared to a popular 3-step BPO based product in adult patients with mild-to-moderate acne vulgaris. Therefore, we conducted a randomized controlled split-face study to investigate the efficacy, safety, and tolerability of both OTC regimens.

## Materials and Methods

### Regimens

Two OTC regimens were compared. The first OTC regimen (Geologie, New York City, New York) included 3 separate products: 1) a cleanser (2% salicylic acid), 2) a day cream (5% azelaic acid, 2% hyaluronic acid, and 1% niacinamide), and 3) a night cream with graduated levels of retinol to maximize patient tolerability. From baseline to week 4, subjects were given a night cream with 0.1% retinol. From week 4 to week 8, subjects were given a night cream with 0.2% retinol. Finally, from week 8 to week 12, subjects were given a night cream with 0.3% retinol. The second OTC Regimen (Proactiv Solution, Southaven, Mississippi) also included 3 separate products: 1) a cleanser (2.5% BPO), 2) a toner (glycolic acid), and 3) repairing treatment (2.5% BPO). Proactiv Solution was selected given its high level of popularity as a BPO routine, and a similar 3-step routine as the Geologie Clear System. All products from each OTC regimen were transferred to unlabeled bottles to maximize blinding. Instructions on applications and use were provided per each manufacturers’ instructions. The dermatologist performing skin lesion grading was blinded to treatment laterality.

### Study Design, Enrollment, Inclusion / Exclusion Criteria, and Study Endpoints

This was a prospective, randomized controlled, double-blinded, single-center study (clinicaltrials.gov: NCT05446402) conducted at Northwestern University in the Department of Dermatology after IRB approval (STU00217056). Patients were recruited in person and via social media (e.g., Facebook). A split-face design was used to compare the Geologie Clear System versus the BPO routine. All eligible subjects (>18 years of age with a clinical diagnosis of mild or moderate acne vulgaris and without an active skin infection or known allergy to the ingredients being evaluated) were recruited and consented. Acne lesion count (open comedones, closed comedones, papules, pustules, nodules, and cysts) was determined by a blinded board-certified dermatologist (PV) at baseline, Week 4, Week 8, and Week 12 of the study via the Facial Lesion Count.^7^ Patients self-collected images with standard mobile phones. Subjects were asked to complete both tolerance and product preference surveys at Week 4, Week 8, and Week 12. For tolerability, subjects were asked to rate their level of redness, itching, and burning on a 5-point scale with 1 being very mild and 5 being very severe. For patient preferences, subjects were asked to complete questions related to the product’s effect on their skin, and their propensity to use the product in the future on a 5-point scale. The primary endpoint was acne lesion count with secondary endpoints including both patient preference and tolerability survey results.

As a non-inferiority study, 20 subjects would enable the detection of an absolute difference of 20% with at least 80% power and a 5% level of significance if the standard deviation of this difference is no larger than 50%. Adjusting for a drop rate of 25%, a target of 27 subjects for recruitment and consent was set. The data were analyzed using Stata. Aggregated data for lesion counts were determined for each time point during the study (baseline, Weeks 4, 8, and 12 of treatment use). Descriptive statistics are presented as means. Paired t-tests were used to compare performance with all p-values were two-tailed assuming equal variances with a level of significant of p_≤_0.05.

## Results

A total of 37 subjects were recruited and consented for the 12-week study. Expansion of the initial target recruitment was required due to drop out of three subjects due to intolerance of the BPO routine and a higher than anticipated lost-to-follow up rate (n=13) (Figure 1). Prior to starting both regimens, the final cohort of patients (n=21) completed a 2-week washout period where no acne treatments, prescription, or OTC medications were used. The final analysis cohort included 10 females, 10 males, and 1 not reported (Table 1). Most subjects were between the ages of 20-29 (n=10) and 30-39 (n=6). Self-reported skin type was most reported as a combination of oily and dry (61%; n=13/21)

**Table 1.** Final demographics of n=21 subjects with mild-to-moderate acne vulgaris.

| Demographics |  |
| --- | --- |
| Total | 21 |
| Sex |  |
| Male | 10 |
| Female | 10 |
| Prefer Not to Say | 1 |
| Age |  |
| 20-29 | 10 |
| 30-39 | 6 |
| 40-49 | 2 |
| Not Reported | 3 |
| Skin Type |  |
| Dry | 2 |
| Normal | 2 |
| Combination | 13 |
| Oily | 4 |
| Embarrassment from Skin |  |
| Very Much | 1 |
| A Lot | 4 |
| A Little | 13 |
| Not at All | 3 |
| Ethnicity |  |
| Asian | 5 |
| Black | 2 |
| Hispanic or Latino | 4 |
| White | 8 |
| Other | 2 |

**Figure 1.**
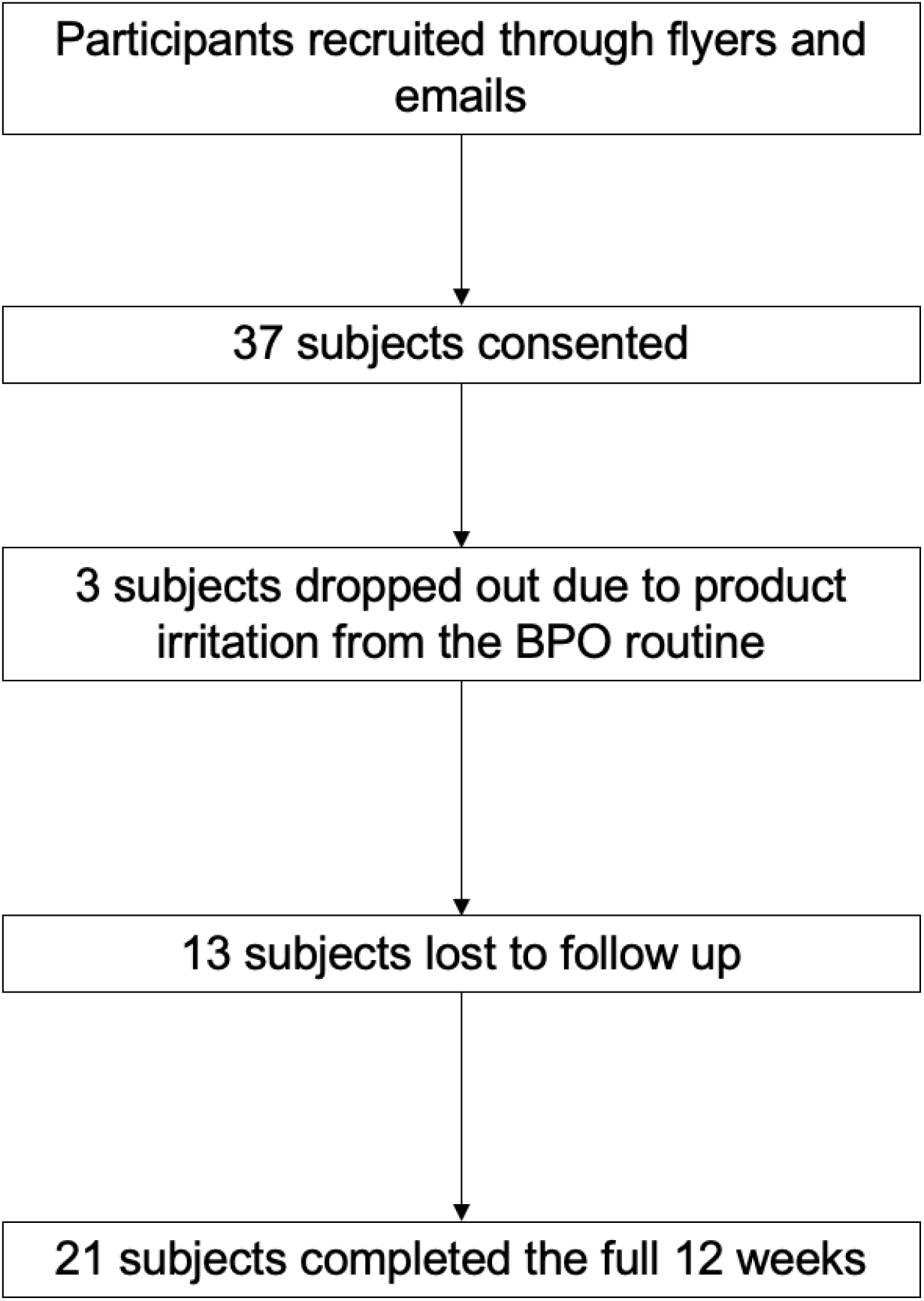
Flow chart of recruitment and final analysis dataset.

Both Geologie and the BPO routine demonstrated a high degree of efficacy evidenced by reduction in facial lesions. At week 4, both regimens had a similar mean number of acne lesions (Geologie: 7.2 and Proactiv: 7.5) with no statistical difference (p=0.80). At week 8, Geologie had an average of 6.6 acne lesions and the BPO routine had an average of 6.5 acne lesions (p=0.94). At week 12, Geologie had an average of 4.6 acne lesions and the BPO routine had an average of 4.7 acne lesions (p=0.93). Over the entire 12 weeks, each product reduced total acne lesions by 36% (Geologie Clear System) and 40% (BPO routine).

Detailed patient preference and user tolerability in Week 4 are shown in Figure 2. Across a total of 25 domains, the Geologie Clear System outperformed the BPO routine in 19 domains (76%). Geologie’s highest performing categories were a patient’s likelihood to use the product in the future, skin feel, and preference. In addition, the Geologie Clear System was preferred in domains for efficacy (faster acne clearing, skin less oily) and skin look and feel (softer, smoother, brighter, and more hydrated). The BPO routine was favored in other categories related to acne prevention. When aggregating both user preferences and tolerability Geologie was more preferred than the BPO routine in 79% of domains (22/28) at Week 4. In Figure 3, users reported higher scores for the Geologie Clear System in skin feel, and future product usage at Week 4 and Week 8. At Week 12, these differences were less apparent (Figure 3).

**Figure 2.**
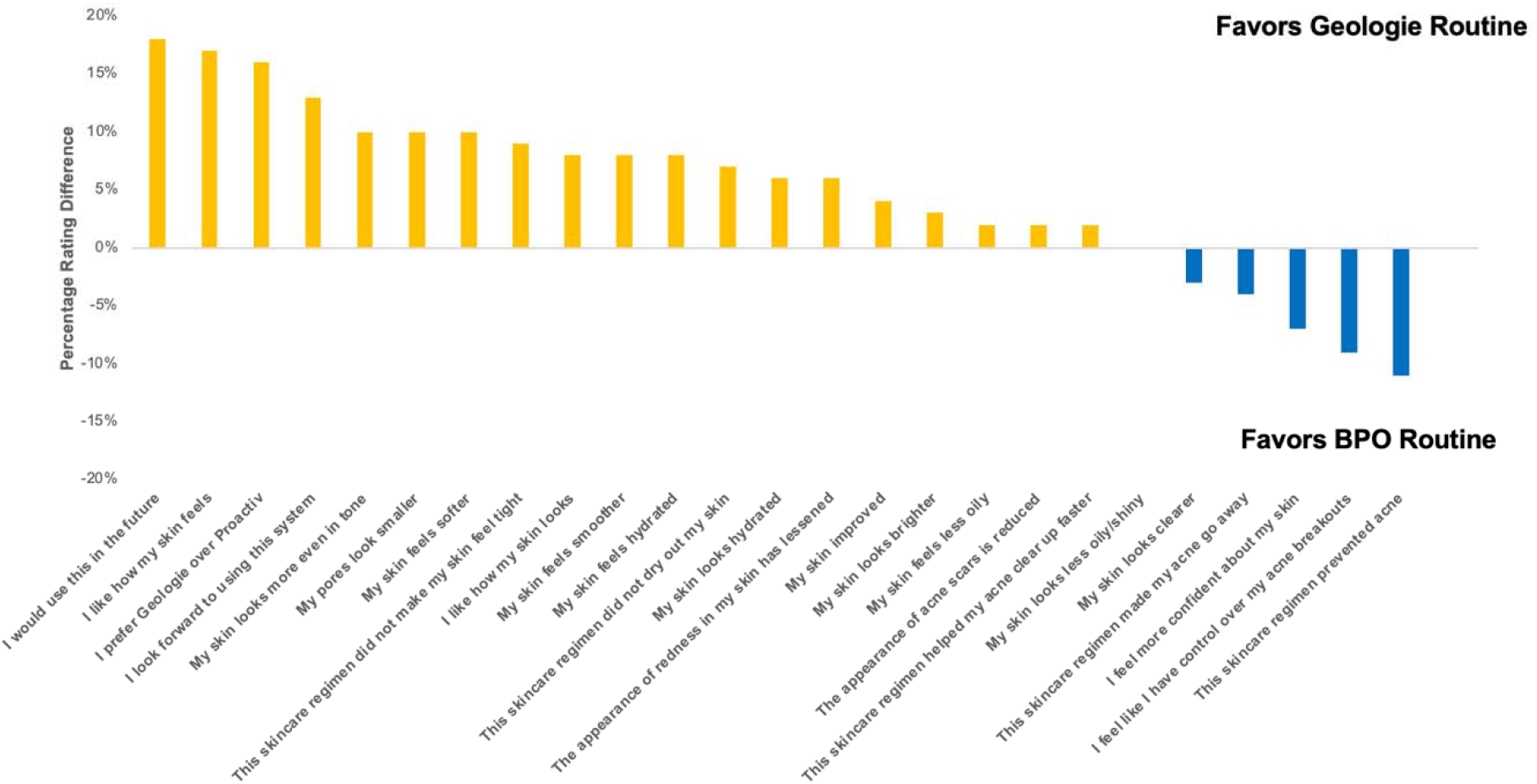
Detailed tolerability and preference survey conducted at Week 4 showed greater preference for the Geologie product across the majority of domains.

**Figure 3.**
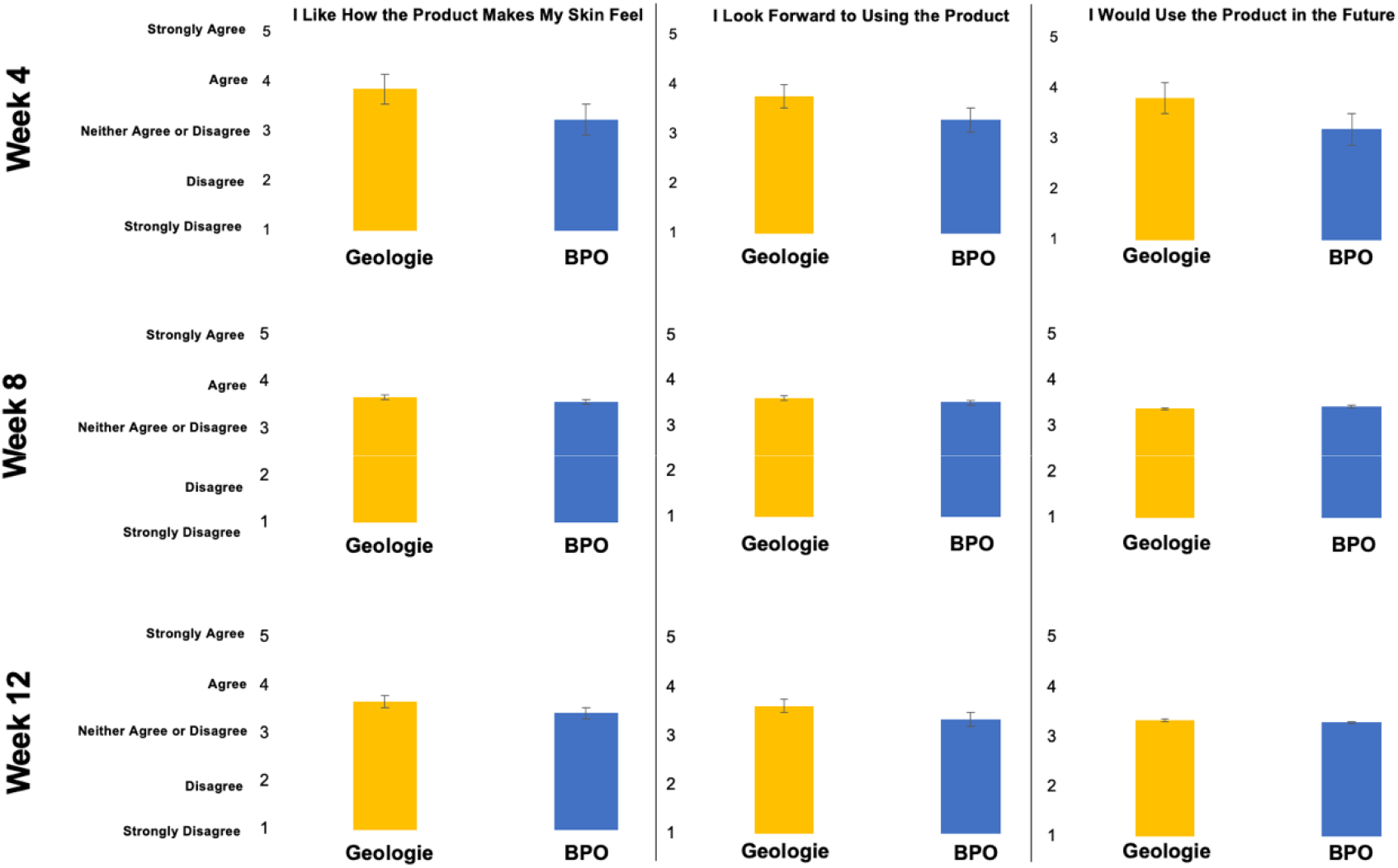
Week 4, Week 8, and Week 12 preference survey comparing key domains for the Geologie routine versus the BPO routine.

Additional survey results on tolerability and product preferences were collected at Week 4, Week 8, and Week 12. Detailed Week 4 survey results showed that across 25 domains of user-assessed product performance, In terms of tolerability, Geologie users reported less facial redness, itching, burning, and dryness (Figure 4).

**Figure 4.**
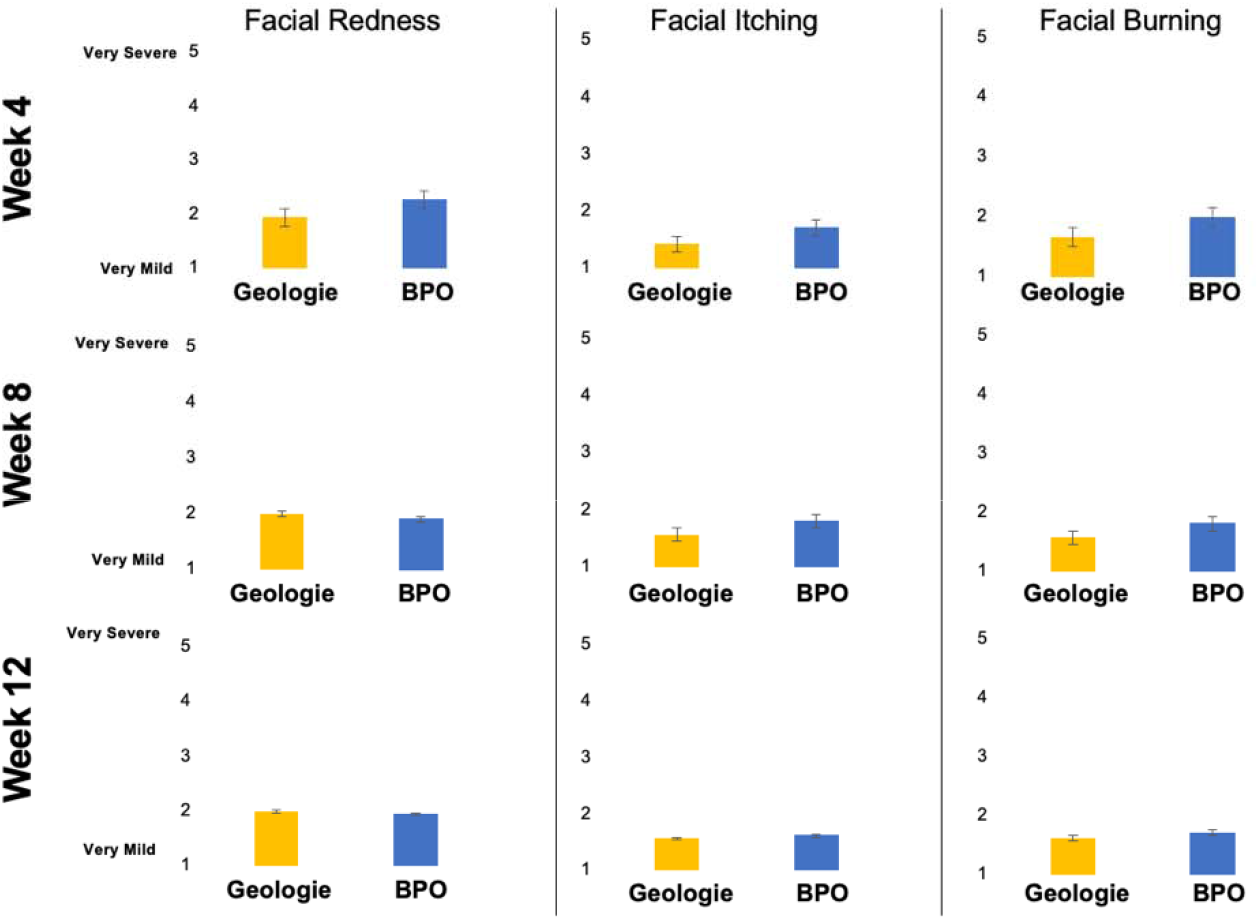
Week 4, Week 8, and Week 12 tolerability survey comparing key domains for Geologie routine versus the BPO routine. Higher preferences were reported for Geologie in Week 4 compared to Week 8 and Week 12.

**Figure 5.**
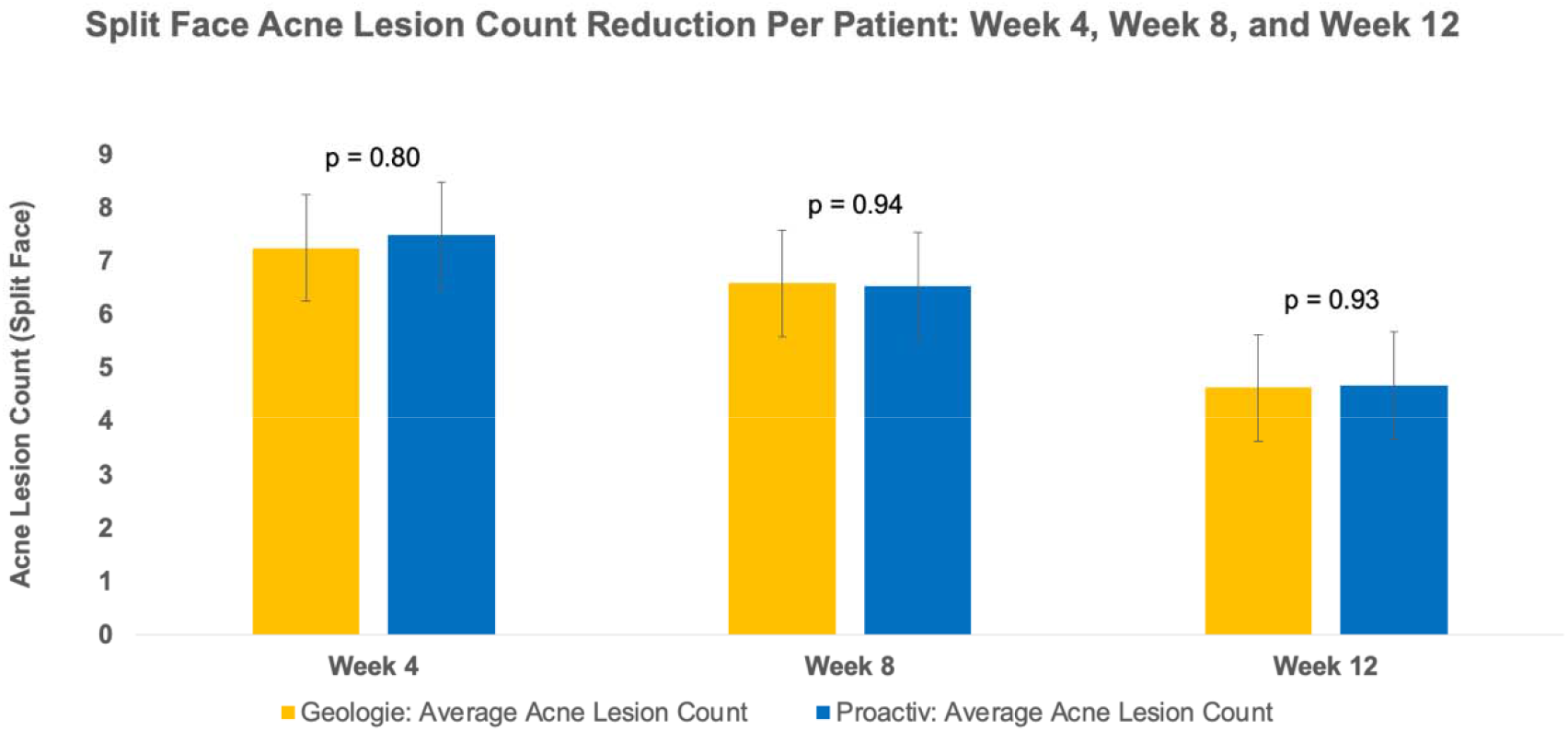
Week 4, Week 8, and Week 12 objective acne lesion (both inflammatory and non-inflammatory) counts for each regimen. There were no statistically significant differences at any week between the Geologie routine versus the BPO routine. The final reduction in objective acne lesions from baseline was 36% for Geologie and 40% for the BPO routine.

### Safety

No adverse events were reported during this study, although three subjects dropped out due to intolerance to the BPO routine during the course of the 12-week study.

## Discussion

The pathogenesis of acne vulgaris is multi-factorial driven by increased sebum production via hyperplastic sebaceous glands, bacterial colonization by *p. acnes*, follicular hyperkeratinization, and inflammation.^8^ BPO based treatments have long been a cornerstone treatment forf acne vulgaris given its ability to reduce antibacterial activity against *Cutibacterium acnes* and suppress sebum production.^9^ However, skin irritation and tolerability remain a key drawback of BPO-based treatments, particularly at higher concentrations^10^—an issue likely exacerbated in the adult acne population with a high underlying prevalence of skin sensitivity. The Proactiv Solution 3-step regimen includes 2 products with BPO and glycolic acid toner. For BPO, the predominate mechanism of action is antibacterial.^11^ Glyolic acid has largely anti-hyperkeratinization activity.^12^ The Geologie regimen was rationally designed to deliver comparable efficacy to BPO based treatments with greater tolerability and less irritation to maximize compliance—this is evident in that no participants dropped out of the study due to intolerance to the Geologie regimen compared to 3 subjects who dropped out due to the BPO routine.

Azelaic acid, already FDA-cleared as a topical treatment for acne in a 20% cream formulation, offers multiple effect anti-acne benefits including being bactericidal for *C. acnes*, anti-inflammatory, and skin lightening for post-inflammatory hyperpigmentation.^13^ Salicylic acid, a beta-hydroxy acid, has also long been a mainstay of acne treatments with a positive effect on abnormal keratinization, and inflammation. Niacinamide also address abnormal sebum production and anti-inflammatory activities. Retinols address hyper-keratinization, provide antibacterial action, and deliver color correction.^12^ Given that retinols, like BPO, have well-established skin irritation and dryness side effects, particularly in treatment naïve or sensitive skin patients, the graduated retinol percentage process (0.1% at month 1, 0.2% month 2, and 0.3% at month 3) was designed to enhance patient tolerability without loss of efficacy. Combination therapies, such as those offered by Geologie, have shown to offer better overall performance than monotherapy strategies.^14^

The overall results show nearly identical efficacy in reducing objective acne lesion reduction at 12 weeks between both Geologie and the BPO routine. In contrast, and particularly at the Week 4 time point, the Geologie regimen exhibited higher ratings by user report across multiple domains from direct product preferences to skin appearance and skin feel. These differences were less apparent at Week 8 and Week 12, although Geologie was still rated higher in skin look and feel, and product perception at the end of the study. In regards to tolerability, Geologie demonstrated less severe redness, itching, and burning with the greatest differences seen at Week 4. These tolerability differences, analogous to the user preference reports, were less evident at Week 12. Overall, tolerability and user acceptance is critical in the management of acne, a chronic disease typically requiring on-going maintenance treatment. User tolerance is particularly relevant among adults with comorbid sensitive skin. Between 30% to 65% of all patients with acne do not adhere to a treatment regimen and as a result 50% do not receive the full benefits of treatment.^15^ For OTC regimens, skin irritation and drying is the most common side effect.^16^ To drive greater adherence and overall treatment success, products should be both effective and also tolerable to use.^14,17^ These results suggest that positive early experiences by new users is especially important—the Geologie system performed better in 79% of user reported domains in efficacy, product performance, and tolerability at the critical 4 week mark. Though outside the scope of this study, early positive experiences may encourage continued and longer-term adherence to treatment reducing rates of drop out among new users and long term benefits in terms of acne reduction.

There are some relevant limitations to note. While a randomized double-blind split-face design lowers the risk of confounders, the final analysis set included only 21 subjects. The high dropout rate where patients were lost to follow up may reduce the confidence in the overall results—we anticipate this to largely be due to high survey burden. Future work should expand on these initial findings in a larger cohort.

## Conclusion

A rationally designed 3-step regimen with azelaic acid, niacinamide, salicylic acid, and graduated nightly retinol resulted in comparable acne lesion reduction with higher overall patient tolerability and patient preferences compared to a 3-step regimen with BPO and glycolic acid in a randomized, double-blind, split-face study of 21 adult patients with acne vulgaris over 12 weeks of therapy.

## Data Availability

All data produced in the present study are available upon reasonable request to the authors

## Acknowledgements

None

## Notes

### Competing Interest Statement

SX has stock options and is a consultant of Regimen - the manufacturer of one of the products studied here. JRW has a spouse with stock options and a consultant of Regimen - the manufacturer of one of the products studied here.

### Clinical Trial

NCT05446402

### Funding Statement

This study was funded by Regimen

### Author Declarations

Ethics committee/IRB of Feinberg School of Medicine gave ethical approval for this work

### Summary of Updates

The title was changed from "Clinical Study" to "Clinical Trial".

